# Saliva Is Comparable to Nasopharyngeal Swabs for Molecular Detection of SARS-CoV-2

**DOI:** 10.1101/2021.04.21.21255621

**Authors:** Cody Callahan, Sarah Ditelberg, Sanjucta Dutta, Nancy Littlehale, Annie Cheng, Kristin Kupczewski, Danielle McVay, Stefan Riedel, James E. Kirby, Ramy Arnaout

**Affiliations:** Department of Radiology; Department of Pathology; Division of Clinical Informatics, Department of Medicine; Beth Israel Deaconess Medical Center, Boston, MA, 02215 USA; Harvard Medical School, Boston, MA, 02215 USA; Abbott Laboratories, Abbott Park, IL, 60064, USA

**Keywords:** SARS-CoV-2, COVID-19, saliva, NP swab, limit of detection

## Abstract

**Background:** The continued need for molecular testing for SARS-CoV-2 and potential for self-collected saliva as an alternative to nasopharyngeal (NP) swabs for sample acquisition led us to compare saliva to NP swabs in an outpatient setting, without restrictions to avoid food, drink, smoking, or tooth-brushing.

**Methods:** A total of 385 pairs of NP and saliva specimens were obtained, the majority from individuals presenting for initial evaluation, and were tested on two high-sensitivity RT-PCR platforms: the Abbott m2000 and Abbott Alinity m (both with limits of detection [LoD] of 100 copies of viral RNA/mL).

**Results:** Concordance between saliva and NP was excellent overall (Cohen’s κ=0.93), for both initial and followup testing, for both platforms, and for specimens treated with guanidinium transport medium as preservative as well as for untreated saliva (κ=0.88-0.95). Viral loads were on average 16x higher in NP specimens than saliva specimens, suggesting that only the relatively small fraction of outpatients (∼8% in this study) who present with very low viral loads (<1,600 copies/mL from NP swabs) would be missed by testing saliva instead of NP swabs, when using sensitive testing platforms. Special attention was necessary to ensure leak-resistant specimen collection and transport.

**Conclusions:** The advantages of self-collection of saliva, without behavioral restrictions, will likely outweigh a minor potential decrease in clinical sensitivity in individuals less likely to pose an infectious risk to others for many real-world scenarios, especially for initial testing.

**Key points:** Saliva has comparable sensitivity and specificity to nasopharyngeal swabs for RT-PCR-based COVID-19 testing (concordance, κ=0.93; *n*=385 participants), albeit with slightly lower recovery of viral RNA. Treatment with a readily available guanidinium preservative within 24 hours of sample collection improves recovery.

## Introduction

The currently accepted gold-standard method for diagnosing infection with SARS-CoV-2 is performing RT-PCR on nasopharyngeal (NP) secretions collected using an NP swab. This is choice is consistent with and was in part based on experience with other respiratory pathogens such as influenza viruses. However, this nasopharynx has the drawback of requiring experienced healthcare staff to perform sample collection, and such personnel are in chronic short supply. Also, requiring potentially infectious patients to congregate at a collection site is not ideal as it may inadvertently lead to iatrogenic infection. These suboptimalities have led to a number of investigations into whether and under what clinical circumstances alternative specimen types might be able to substitute for NP swabs [1–12]. Nasal secretions have the advantage of being able to be self-collected by swabbing the anterior nares; however, nasal secretions been shown to be less sensitive than NP secretions for viral loads below ∼1,000 copies/mL [5,6,13] and still require a swab for collection (swabs have been in intermittently short supply over the course of the pandemic) [14].

Saliva has the advantage of requiring neither trained personnel nor a swab, making it an attractive alternative to both NP- and/or nasal-swab testing. Saliva can be self-collected using only a sterile container, making it amenable to at-home/off-site collection. Sensitive detection of respiratory viruses from saliva had been shown prior to the pandemic. [15,16]. Consequently, saliva has been the subject of a number of studies in the context of the pandemic; however, these have reached varying conclusions regarding the suitability of saliva as an alternative to NP swabs, with some studies showing complete concordance and others only moderate agreement [9]. Known and potential reasons for these differences include timing/acuity of patients’ clinical presentation, storage/processing conditions of collected samples, and sensitivity of the respective testing platforms used for detection of SARS-CoV-2. To address these issues, we compared saliva to NP swabs in 1,600 subjects across two testing platforms (Abbott m2000 and Abbott Alinity m) and two sample-processing approaches (untreated/”neat” and treated with guanidinium as a preservative) for individuals presenting for initial presentation vs. followup testing.

## Methods

### Institutional review

This study was reviewed and approved by institutional review board at the Beth Israel Deaconess Medical Center (BIDMC; IRB protocol no. 2020P000769).

### Trial design

This was a multi-arm study: initial vs. followup presentation, two different testing platforms (Abbott m2000 and Abbott Alinity m), and two different sample-processing pipelines for the saliva samples (unadulterated/”neat” saliva vs. preserved (“treated”) with guanidinium isothiocyanate [GITC], i.e. Abbott multi-collect transport media, part of the Abbott Multi-Collect Specimen Collection Kit, catalog no. 09K12-004; Abbott Laboratories, Abbott Park, IL). See Fig. 1.

**Figure 1.**
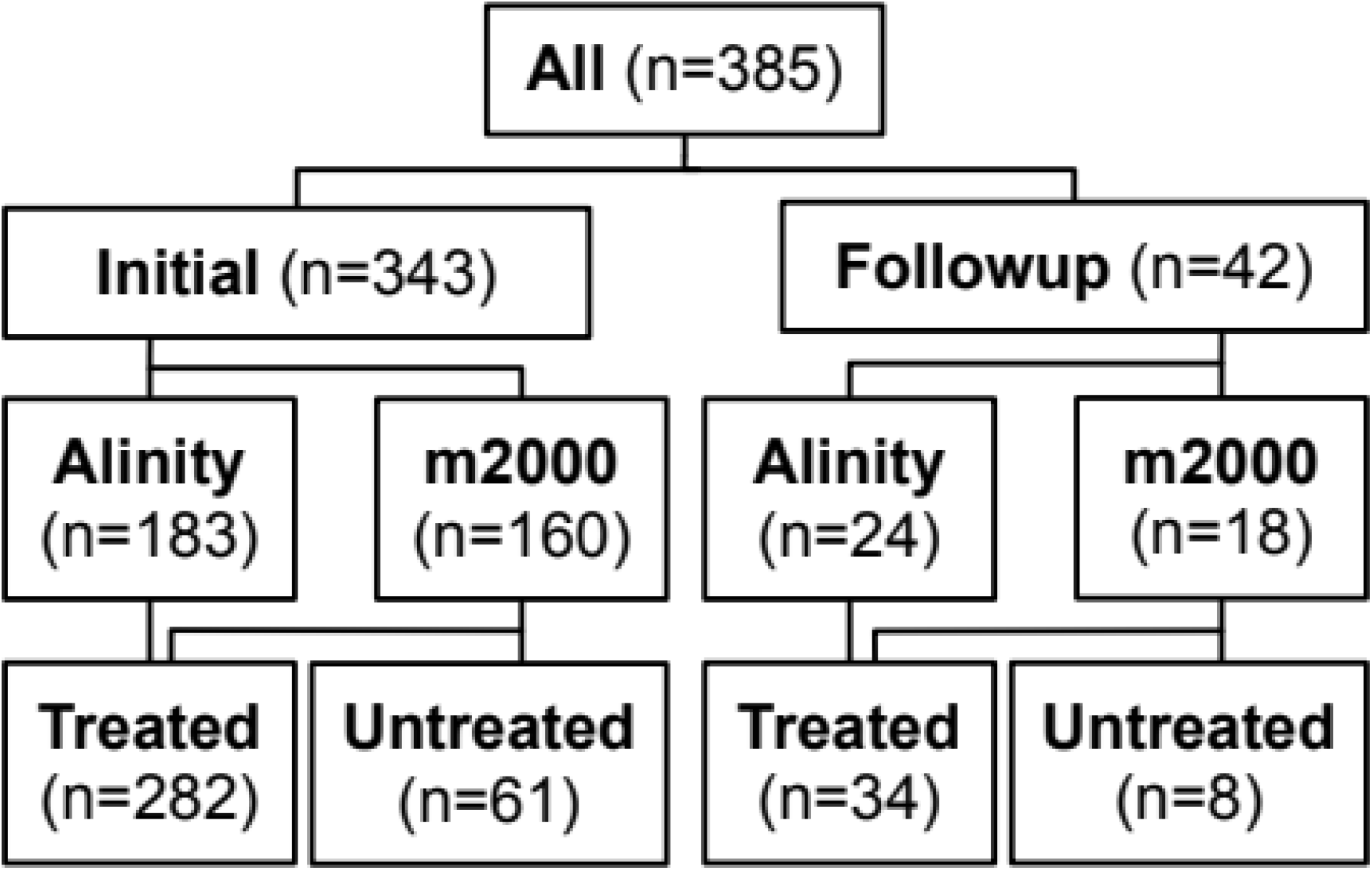
Numbers of participants in each study arm.

### Trial participants and sample collection

Informed consent was obtained from English-speaking adults presenting for either initial or followup testing for COVID-19 at BIDMC and Beth Israel Deaconess Chelsea drive-through collection sites.

#### Saliva specimens

While waiting in line for NP testing, each participant was given a sterile sample collection cup (VWR International, part no. 76299-868) and asked to spit/drool to a clearly labeled 3mL fill line. No exclusions were made on the basis of recent consumption of food or drink or of smoking, and participants were not asked to rinse the mouth or perform any other preparation ahead of sample collection. They were informed that 3mL is a large amount of saliva and may take several minutes/deposits to achieve, and told that thinking of a favorite food may help trigger salivation. They were told to close the cup tightly when finished and to hand it to the healthcare worker who would be acquiring the NP specimen.

#### NP specimens

NP specimens were collected per standard protocol. Briefly, the NP swab was inserted into the nasopharynx and rotated for 10 seconds, removed, and placed in a vial containing viral transport media [17].

*Transport*. Saliva specimens were individually bagged and transported together with the NP specimen via courier to BIDMC’s central laboratory for testing. Courier frequency was every 1-2.5 hours, with the maximum transport time to the laboratory being approximately 20 minutes. Specimens were transported at room temperature. At the BIDMC Pathology central laboratory, specimens were logged in and processed for testing as described below.

### Sample processing

For GITC-treated saliva samples, 1mL of saliva was added to the multi-Collect tube that contains 1.25mL GITC buffer (“treated”). For untreated saliva samples, 1mL was used “neat.” Samples with <1mL were excluded.

At the start of the study, treated, untreated, and a 1mL aliquot of NP sample from the same participant were briefly vortexed (2-5 seconds) and amplified using Abbott m2000 RealTime

SARS-CoV-2 Assay on an Abbott m2000 RealTime System or the Abbott Alinity m system. After 16 positive untreated specimens were analyzed on the m2000, the untreated arm was abandoned in favor of the treated arm. Thereafter, treated specimens were stored at 4°C until the paired NP sample had resulted. If the NP sample was positive, the GITC sample was run alongside an additional 1mL aliquot from the NP sample. Categorical (positive vs. negative), Ct, and IC values were recorded. Cutoff for positive was a Ct value of 31.5 on the m2000 and 42 on the Alinity m.

### Limits of detection (LoD)

*NP*. The LoD for SARS-CoV-2 from NP swabs was validated at 100 copies/mL on both the m2000 and Alinity m platforms, as described previously [18].

#### Saliva

To determine the LOD for saliva, a (non-infectious) recombinant, enveloped, positive-single-stranded RNA virus containing SARS-CoV-2 RNA (AccuPlex COVID-19, Seracare, Milford, MA, USA; 1.3×10^7^ copies/mL as determined by digital PCR) was initially diluted into RNA Storage Solution, purchased from Invitrogen (AM7001; ThermoFisher Scientific, Waltham, MA, USA) to make a positive stock. The positive stock was then spiked to target concentrations in pooled negative saliva prepared with purchased leftover remnant deidentified specimens. A portion of these spiked “neat” or “untreated” samples were combined with transport medium using a saliva: GITC volume ratio of 1:1.25; these are “treated” samples. Tested concentrations were 900, 675, 450, 225, and 112.5 copies/mL for untreated samples and 400, 300, 200, 100, and 50 copies/mL for treated samples. The initial LoD was determined by testing targeted levels in replicates of 3; the final LoD was confirmed by testing in 21 additional replicates of the lowest two dilutions.

### Stability testing

Saliva specimens were collected as described above and stored at 4°C pending results of the paired NP specimen. Twenty NP-confirmed negative saliva specimens were pooled. The pooled sample was spiked to a final concentration of 11,215 copies/mL of SeraCare material and aliquotted into 24 × 1.1mL replicates. Starting immediately (the 0-hour timepoint), each of three replicates was treated with 1.2mL GITC and promptly run on the Alinity m platform per standard protocol. This was repeated at 4, 8, 12, 16, 20, 24, and 45 hours.

Replicates were stored at room temperature between time points. Categorical, Ct, and IC values were recorded.

### Sample-processing times

NP specimens were tracked using BIDMC’s clinical data system, with the following recorded: order creation time as a surrogate for sample collection time (±15 minutes), lab-control time (sample login at BIDMC), login time at BIDMC’s molecular diagnostics laboratory, archive time, and result time.

### Statistical analyses

For concordance testing, RT-PCR results were considered categorically as either positive or negative; testing agreement was assessed by Cohen’s kappa (*κ*) [19].

Conversion to viral load was performed as described previously [20]. “Initial testing” was defined as testing of a saliva sample collected fewer than five days after the first COVID-19 RT-PCR test. “Followup testing” was defined as testing occurring thereafter. Leaking samples and internal-control failures were omitted from analysis.

### Significance testing

We tested whether Ct values for a given subset of saliva samples differed from the Ct values for the paired NP swabs (the controls) using Wilcoxon’s signed rank test. This tested the null hypothesis that values for controls and prototypes are drawn from the same underlying distribution. The Benjamini-Hochberg approach [21] was used to control the false-discovery rate (FDR) at 1%.

### Software

We used Python (v3.6-3.8) and its NumPy, SciPy, Matplotlib, Pandas, and ct2vl libraries for the above analyses and related visualizations.

### Data Availability

De-identified data is presented in Supplementary File 1. Supplementary Data can be found at (https://github.com/rarnaout/Covid_diagnostics).

## Results

### Study design and overall observations

Figure 1 depicts the design of this study of adults being tested by RT-PCR for infection with SARS-CoV-2 and the numbers of participants tested in each arm. Participants waiting for NP sample collection were asked to spit into a sterile urine cup clearly marked with a 3-mL fill line until the line was reached. Participants were informed that this would likely require spitting more than once, encouraged to persist despite any embarrassment or distaste/mild disgust, and instructed to thread the lids carefully and close them tightly to avoid leaking. Study staff retightened cups as necessary. Saliva specimens were double bagged and shipped alongside the NP specimen. Overall, this limited the incidence of leaking saliva, which was encountered frequently initially.

### Limit of detection of SARS-CoV-2 in saliva on Abbott m2000 and Alinity m

In spiking experiments, the lowest concentration level with observed positive rates ≥95% was 100 copies/mL in treated saliva and 225 copies/mL in untreated saliva. Logistic regression estimated LoDs of 45 copies/mL in treated and 104 copies/mL in untreated saliva (76 copies/mL combined), statistically comparable to the previously validated LoD for NP swabs on these platforms, 100 copies/mL (Fig. 2). (Note that treatment results in a roughly 2-fold dilution of the sample.)

**Figure 2.**
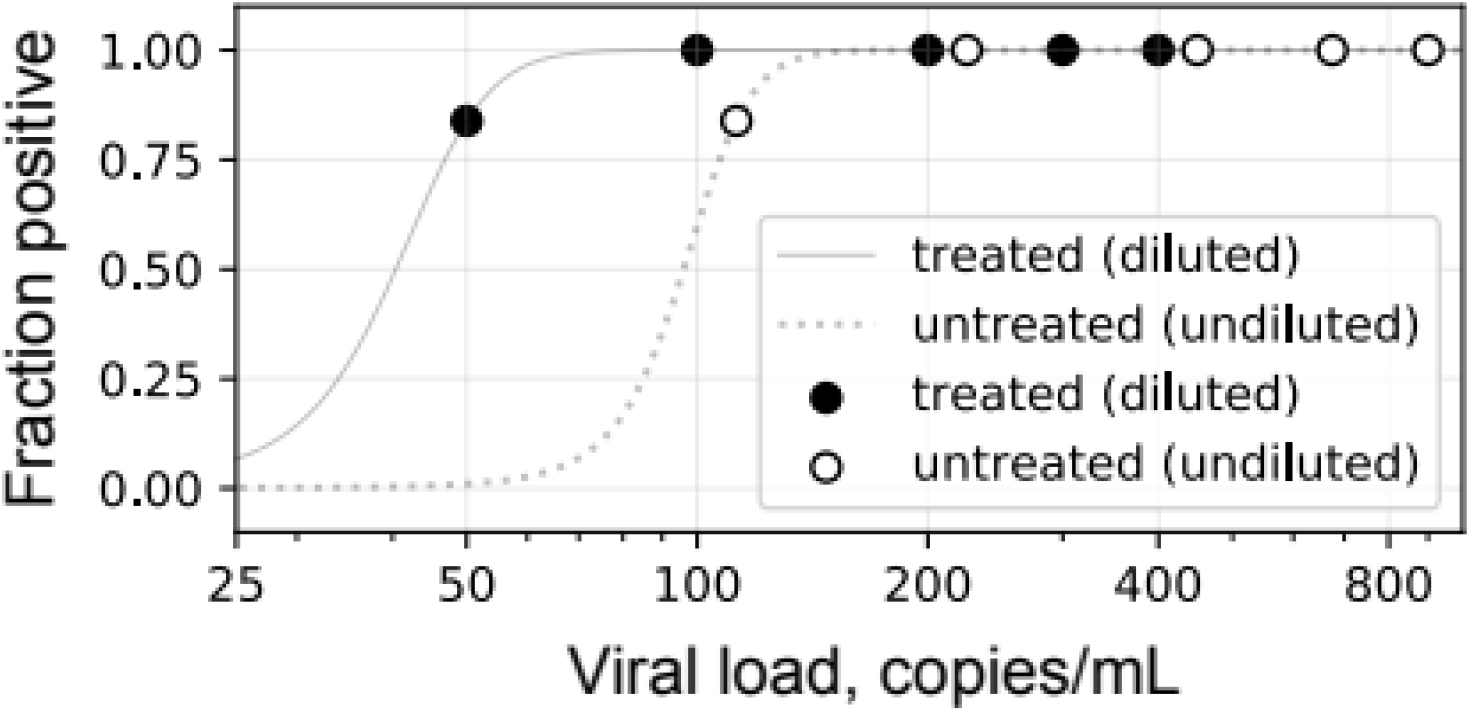
LoD for treated and untreated saliva on Abbott m2000 and Alinity m platforms. Lowest two datapoints, 24 replicates each; others in triplicate.

#### Stability of SARS-CoV-2 in saliva over time

Replication-incompetent, enveloped, positive single-stranded RNA Sindbis virus into which SARS-CoV-2 genomic material was cloned as a non-infectious SARS-CoV-2 surrogate was stable in untreated saliva at room temperature, with concentration falling by ∼3 fold within the first four hours but then staying stable at that level through two days (Fig. 3). Saliva treated with the preservative GITC was stable for at least 24 hours. Together, these findings support the viability of workflows in which, as a limiting timeline, saliva is collected and transported “neat” and treated with GITC within two days for subsequent testing.

**Figure 3.**
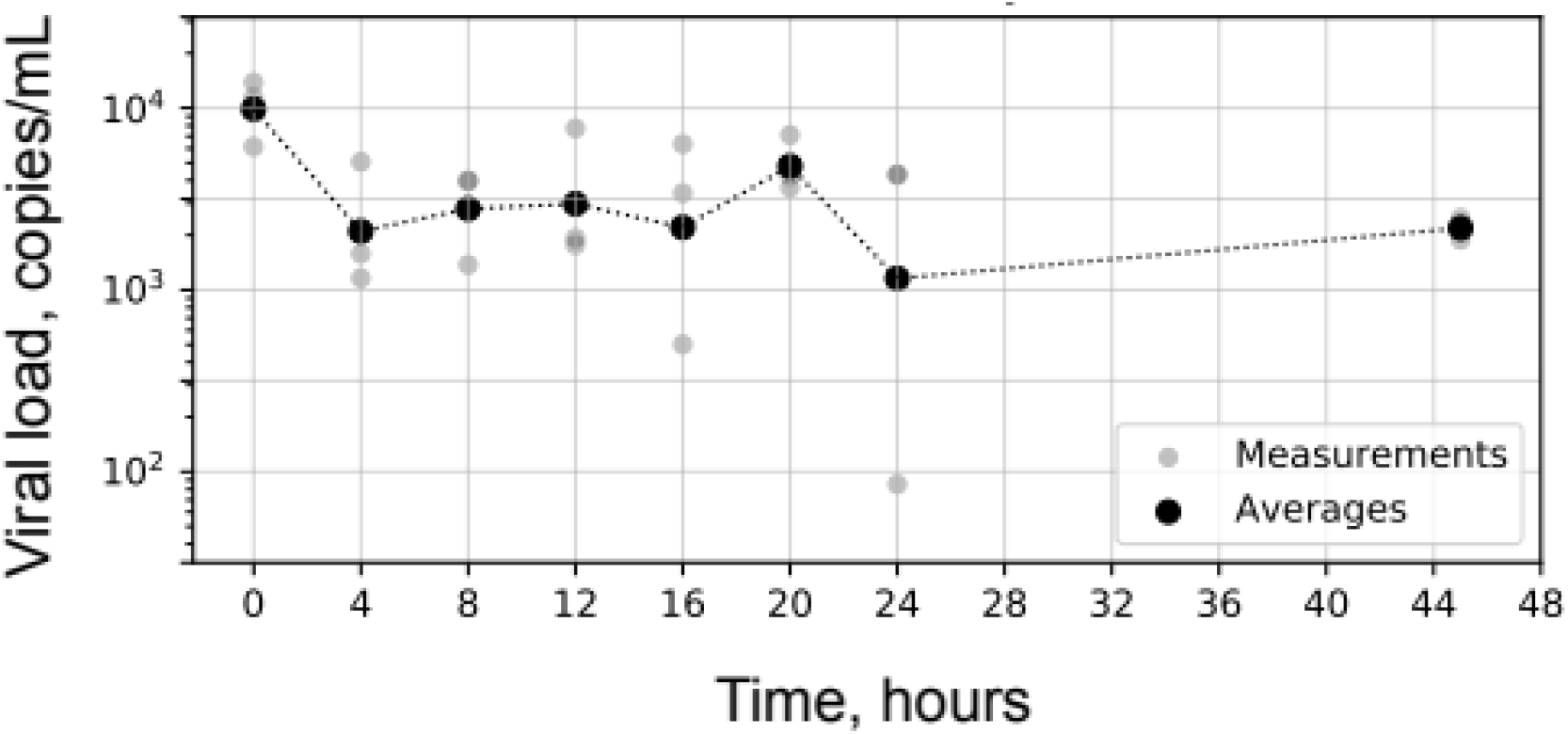
Stability of replication-incompetent SARS-CoV-2 surrogate virus in untreated saliva samples over time. Triplicate measurements in gray; geometric means in black.

### Comparison of saliva to NP swabs

Overall, saliva results were highly concordant (Cohen’s κ=0.93) with results from paired NP swabs obtained from participants in a medium-to-high prevalence population(∼20% positivity) who presented for screening and follow-up, as tested on two sensitive RT-PCR platforms, the Abbott Alinity m and Abbott m2000 (LoD 100 copies/mL) (Fig. 4). Using the LoD as the cutoff, there were only nine discordant results among 385 paired samples, and these were evenly balanced (4 NP-swab-positive/saliva-negative samples vs. 5 saliva-positive/NP-swab-negative samples). Despite the high concordance, (geometric) mean viral load in NP samples was ∼16 times as high as in saliva samples (3.1 million vs. 200,000 copies/mL, respectively; Wilcoxon *p* value for difference of saliva vs. NP viral load distributions, 2×10^−6^). These trends were robust across all subgroups analyzed, whether by initial testing (*n*=343, κ=0.93) or follow-up testing (*n*=42, κ=0.88); whether samples were treated with GITC as soon as they reached the laboratory (Fig. 5) (*n*=316, κ=0.92) or not (n=69, κ=0.95); and whether they were run on the Alinity m (n=207, κ=0.91) or m2000 (n=178, κ=0.94). Notably, mean viral load for NP-swab samples was 13 times as high as for saliva samples treated with GITC vs. 40 times as high for untreated saliva samples. Mean viral load was 2-3x as high for participants presenting for their initial COVID-19 test vs. follow-up (3.7 million vs. 1.5 million copies/mL, respectively, as measured on NP samples, and 240,000 vs. 81,000 copies/mL, respectively, as measured on saliva samples).

**Figure 4.**
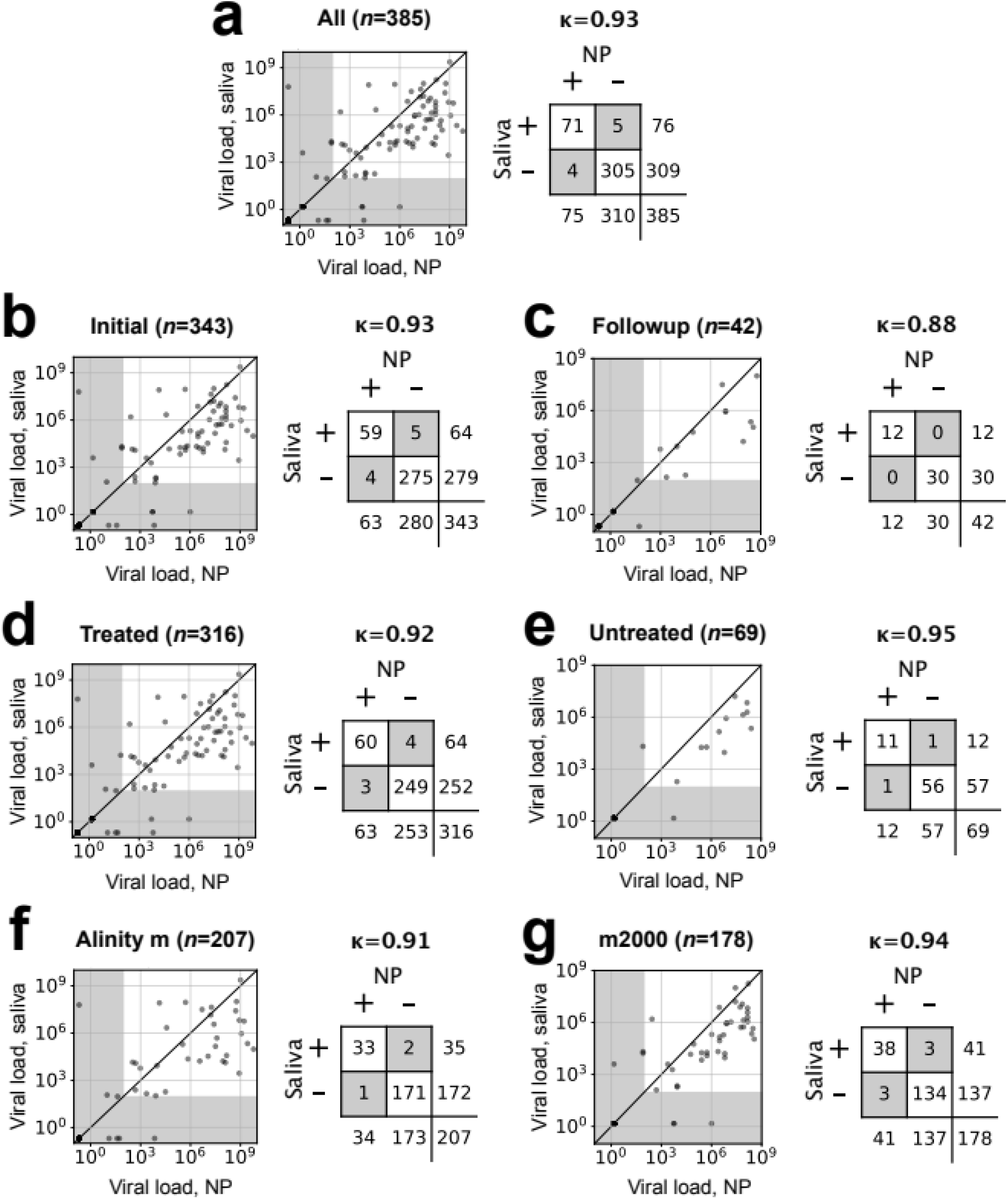
Viral load in saliva vs. NP swab samples, Cohen’s kappa (κ) concordance values, and contingency tables for (a) overall study, (b) subjects presenting for initial presentation (within 5 days of first COVID-19 RT-PCR test), (c) subjects presenting for followup testing, (d) samples treated with GITC transport buffer as a preservative after receipt at the central laboratory, (e) untreated samples, (f) samples run on the Alinity m platform, and (g) samples run on the m2000. Diagonal lines in scatterplots, 1:1. Gray shaded areas in the scatterplots are below the LoD (100 copies/mL). Gray shaded cells in the contingency tables highlight discordant results.

**Figure 5.**
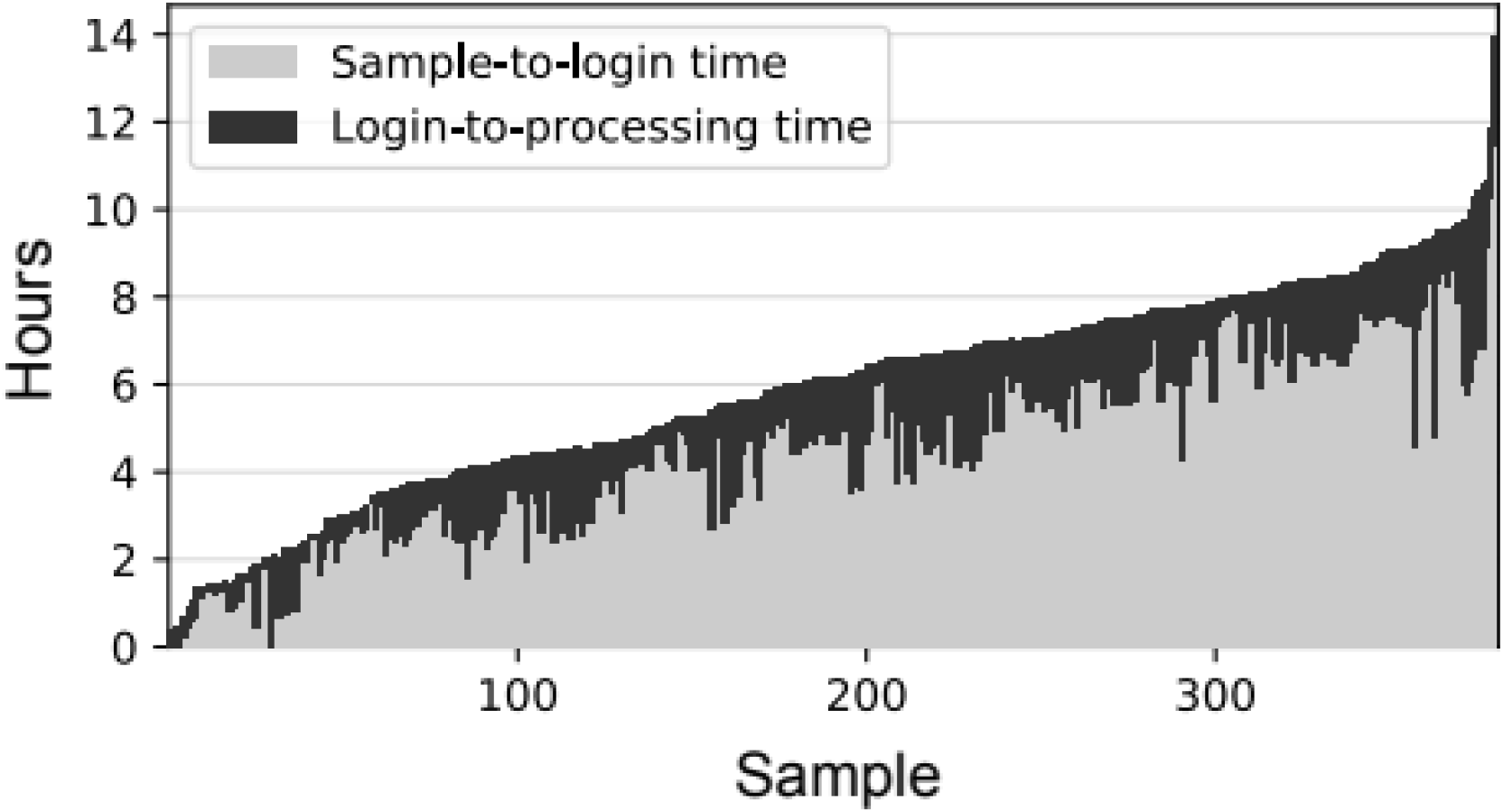
Sample processing times, ranked by increasing order of total time. Sample-to-login time is the time between sample acquisition and login at the central laboratory. Login-to-processing time is the time between login and arrival of the specimen in the molecular laboratory where GITC treatment and RT-PCR testing are performed.

## Discussion

While saliva testing has shown promise as a suitable specimen for detection of several viral respiratory pathogens [8–10,15], NP swabs remain the gold standard in most clinical settings, requiring well-trained healthcare staff to obtain them. The demands of the COVID-19 pandemic have led to a number of investigations of saliva for molecular detection of SARS-CoV-2 in a wide range of clinical and non-clinical settings and with a variety of patient instructions and processing steps, with the overall conclusion that saliva is comparable to but slightly inferior to NP swabs [22]. Some studies have instructed patients to provide morning specimens or to abstain from eating/drinking/smoking before providing a specimen, constraints that complicate collection. We sought to test how well saliva testing in a real-world outpatient setting without any such constraints would perform, with the only special handling step being adding a preservative, in the form of GITC transport buffer once the specimen arrived from the outpatient site to our hospital’s central laboratory for testing.

The overall results in our current study were consistent with previous work [22], with very high concordance between saliva and NP swabs under all scenarios tested. A minor but surprisingly important observation was the importance of leak-resistant collection cups, as a substantial number of collected specimens were unusable due to leaks, despite proper instruction for sample collection. The leak-resistant cups we adopted mitigated leaking somewhat, as they incorporated a soft gasket-like seal inside the lid, with a thread length to pitch ratio of ∼3. Cups using a more deformable gasket or larger ratio may further reduce sample leakage.

Nevertheless, our experience demonstrates that simple and readily available specimen-collection cups can be repurposed for saliva collection for respiratory-virus testing. Usefully, we observed high concordance without any restrictions on participant activities (eating/drinking/smoking/tooth brushing; the color and heterogeneity of several saliva samples was highly suggestive of these activities); without asking participants to rinse, cough, or retch; and without requiring specimens to be obtained in the morning (indeed most of samples were collected in the early afternoon). While it cannot be ruled out that these additional requirements might improve performance (∼5% improvement in concordance in meta-analysis (20)), the results of the present study suggest that the procedural hurdles of obtaining specimens with these constraints likely outweigh any modest incremental benefit.

Furthermore, we demonstrated that SARS-CoV-2 viral material was stable in GITC-treated saliva for at least 24 hours. The time between sample collection and addition of GITC never exceeded 12 hours. Treatment was performed in the laboratory and not earlier, at the collection site, for simplicity of workflow. We chose not to include the preservative in the specimen-collection cups for concern over potential exposure of participants to a caustic substance. The median time between specimen collection and addition of GITC was approximately 6 hours (Fig. 5); however, we demonstrated that viral material is fairly stable in saliva at room temperature without preservative, retaining approximately half of viral load at 24 hours and a third at 45 hours, allowing room-temperature transport and login times of over a day. Consistent with these stability studies, untreated saliva samples showed indistinguishably high concordance with NP swabs compared to GITC-treated saliva samples (Fig. 4d-e), although the mean viral load in untreated samples was approximately 3x lower (also consistent with stability studies).

Despite the high concordance overall (κ=0.93) and for all subgroup analyses (κ=0.88-0.95), including for participants presenting for follow-up as well as during initial presentation, the on-average 13x higher viral load detected by NP swabs vs. paired GITC-treated saliva specimens deserves mention. Provided that viral load (as measured by NP swab) is greater than 13 times the platform’s LoD (100 copies/mL in our study), saliva treated with GITC within 24 hours of collection is highly likely to detect infection. However, viral loads less than this threshold (1,300 copies/mL) may be missed. In our study, approximately 92% of initial presentations were above this threshold. The analogous ratio for untreated saliva was 40x, corresponding to a threshold of 4,000 copies/mL; approximately 90% of initial presentations were above this threshold. We conclude that saliva can be fairly expected to detect over 90% of initial SARS-CoV-2 infections in an outpatient setting. To the extent that more severe disease correlates with higher viral loads, performance in inpatient settings is likely to be even higher. Furthermore, a detection level in saliva that would correspond to 4,000 copies/mL in NP swab samples is likely to capture those who are infectious and is also likely significantly below the limit of detection of currently marketed SARS-CoV-2 antigen tests [20].

In conclusion, we have demonstrated that saliva is comparable to NP-swab collection for molecular detection of SARS-CoV-2 in an outpatient setting, with the disadvantage of slightly lower viral loads. This loss in sensitivity must be weighed against the ease of self-collection. However, we have demonstrated that use of saliva for SARS-CoV-2 molecular detection is both feasible and practical, given suitable specimen-collection containers and transport and processing protocols.

## Acknowledgements

The authors gratefully acknowledge the participation of patients and staff at Beth Israel Deaconess Healthcare | Chelsea for making this study possible. We also acknowledge critical reading of the manuscript, as well as study design input by Joshua Kostera.

## Potential conflicts of interest

This work was supported through a sponsored research agreement with Abbott Molecular. CC, SD, SD, NL, AC, KK, DM, SR, and RA have no other potential conflicts of interest to declare. JK has received reagents from Abbott Molecular for unrelated studies under a Covid-19 Diagnostics Evaluation Agreement.

## References

1. LeBlanc JJ, Heinstein C, MacDonald J, Pettipas J, Hatchette TF, Patriquin G. A combined oropharyngeal/nares swab is a suitable alternative to nasopharyngeal swabs for the detection of SARS-CoV-2. Journal of Clinical Virology 2020; 128:104442.

2. Rhoads DD, Cherian SS, Roman K, Stempak LM, Schmotzer CL, Sadri N. Comparison of Abbott ID Now, Diasorin Simplexa, and CDC FDA EUA methods for the detection of SARS-CoV-2 from nasopharyngeal and nasal swabs from individuals diagnosed with COVID-19. J Clin Microbiol 2020; :JCM.00760-20, jcm;JCM.00760-20v1.

3. McCormick-Baw C, Morgan K, Gaffney D, et al. Saliva as an Alternate Specimen Source for Detection of SARS-CoV-2 in Symptomatic Patients Using Cepheid Xpert Xpress SARS-CoV-2. J Clin Microbiol 2020; :JCM.01109-20, jcm;JCM.01109-20v1.

4. Kojima N, Turner F, Slepnev V, et al. Self-Collected Oral Fluid and Nasal Swabs Demonstrate Comparable Sensitivity to Clinician Collected Nasopharyngeal Swabs for Covid-19 Detection. medRxiv 2020; :2020.04.11.20062372.

5. Berenger BM, Fonseca K, Schneider AR, Hu J, Zelyas N. Sensitivity of Nasopharyngeal, Nasal and Throat Swab for the Detection of SARS-CoV-2. medRxiv 2020; :2020.05.05.20084889.

6. Callahan C, Lee RA, Lee GR, Zulauf K, Kirby JE, Arnaout R. Nasal-Swab Testing Misses Patients with Low SARS-CoV-2 Viral Loads. medRxiv 2020; :2020.06.12.20128736.

7. Azzi L, Carcano G, Gianfagna F, et al. Saliva is a reliable tool to detect SARS-CoV-2. Journal of Infection 2020; 81:e45–e50.

8. Griesemer SB, Van Slyke G, Ehrbar D, et al. Evaluation of specimen types and saliva stabilization solutions for SARS-CoV-2 testing. medRxiv

9. Nagura-Ikeda M, Imai K, Tabata S, et al. Clinical Evaluation of Self-Collected Saliva by Quantitative Reverse Transcription-PCR (RT-qPCR), Direct RT-qPCR, Reverse Transcription–Loop-Mediated Isothermal Amplification, and a Rapid Antigen Test To Diagnose COVID-19. J Clin Microbiol 2020; 58:e01438-20, /jcm/58/9/JCM.01438-20.atom.

10. Ranoa DRE, Holland RL, Alnaji FG, et al. Saliva-Based Molecular Testing for SARS-CoV-2 that Bypasses RNA Extraction. Microbiology, 2020. Available at: http://biorxiv.org/lookup/doi/10.1101/2020.06.18.159434. Accessed 19 March 2021.

11. Teo AKJ, Choudhury Y, Tan IB, et al. Saliva is more sensitive than nasopharyngeal or nasal swabs for diagnosis of asymptomatic and mild COVID-19 infection. Sci Rep 2021; 11:3134.

12. Tu Y, Jennings R, Hart B, et al. Patient-collected tongue, nasal, and mid-turbinate swabs for SARS-CoV-2 yield equivalent sensitivity to health care worker collected nasopharyngeal swabs. Infectious Diseases (except HIV/AIDS), 2020. Available at: http://medrxiv.org/lookup/doi/10.1101/2020.04.01.20050005. Accessed 19 March 2021.

13. Péré H, Podglajen I, Wack M, et al. Nasal Swab Sampling for SARS-CoV-2: a Convenient Alternative in Times of Nasopharyngeal Swab Shortage. J Clin Microbiol 2020; 58:e00721-20, /jcm/58/6/JCM.00721-20.atom.

14. Callahan CJ, Lee R, Zulauf KE, et al. Open Development and Clinical Validation of Multiple 3D-Printed Nasopharyngeal Collection Swabs: Rapid Resolution of a Critical COVID-19 Testing Bottleneck. Journal of Clinical Microbiology 2020; 58. Available at: https://jcm.asm.org/content/58/8/e00876-20. Accessed 17 December 2020.

15. To KKW, Yip CCY, Lai CYW, et al. Saliva as a diagnostic specimen for testing respiratory virus by a point-of-care molecular assay: a diagnostic validity study. Clinical Microbiology and Infection: The Official Publication of the European Society of Clinical Microbiology and Infectious Diseases

16. To KK, Lu L, Yip CC, et al. Additional molecular testing of saliva specimens improves the detection of respiratory viruses. Emerging Microbes & Infections 2017; 6:1–7.

17. Smith KP, Cheng A, Chopelas A, et al. Large-scale, in-house production of viral transport media to support SARS-CoV-2 PCR testing in a multi-hospital healthcare network during the COVID-19 pandemic. Journal of Clinical Microbiology 2020; Available at: https://jcm.asm.org/content/early/2020/05/11/JCM.00913-20. Accessed 10 June 2020.

18. Arnaout R, Lee RA, Lee GR, et al. The Limit of Detection Matters: The Case for Benchmarking Severe Acute Respiratory Syndrome Coronavirus 2 Testing. Clin Infect Dis 2021;

19. McHugh ML. Interrater reliability: the kappa statistic. Biochem Med (Zagreb) 2012; 22:276–282.

20. Arnaout R, Lee RA, Lee GR, et al. The Limit of Detection Matters: The Case for Benchmarking Severe Acute Respiratory Syndrome Coronavirus 2 Testing. Clin Infect Dis 2021;

21. Benjamini Y, Hochberg Y. Controlling the False Discovery Rate: A Practical and Powerful Approach to Multiple Testing. Journal of the Royal Statistical Society: Series B (Methodological) 1995; 57:289–300.

22. Lee RA, Herigon JC, Benedetti A, Pollock NR, Denkinger CM. Performance of Saliva, Oropharyngeal Swabs, and Nasal Swabs for SARS-CoV-2 Molecular Detection: A Systematic Review and Meta-analysis. Journal of Clinical Microbiology 2021; Available at: https://jcm.asm.org/content/early/2021/01/26/JCM.02881-20. Accessed 29 March 2021.

